# Exoskeleton-assisted walking improves pulmonary and exercise performances more than conventional exercise program in individuals with spinal cord injury: a randomised controlled study

**DOI:** 10.1101/2021.10.08.21264727

**Authors:** Xiao-Na Xiang, Li-Ming Zhang, Hui-Yan Zong, Yi Ou, Xi Yu, Yan Liu, Hong-Ying Jiang, Hong Cheng, Hong-Chen He, Cheng-Qi He

**Author notes:** **Corresponding Author:** Hong-Chen He, PhD, Rehabilitation Medicine Centre, West China Hospital, Sichuan University, Chengdu, 610041 Sichuan, PR China., Cheng-qi He, PhD, Rehabilitation Medicine Centre, West China Hospital, Sichuan University, Chengdu, 610041 Sichuan, PR China. These authors contributed equally to this work (Xiao-Na Xiang, Li-Ming Zhang). **Setting:** Department of Rehabilitation Medicine, West China Hospital of Sichuan University.

## Abstract

**Question:** In people with spinal cord injury, does exoskeleton-assisted walking training improve pulmonary ventilation function, motor function and related body structure, walking, and activities of daily life equally comparing those with conventional exercise program?

**Design:** Randomised controlled trial with concealed allocation, assessor blinding and intention-to-treat analysis.

**Intervention:** Both groups undertook 16 sessions of 50-60min training (4 days/week, 4 weeks). Participants in the experimental group received EAW trainings using AIDER system, which assisted standing, walking, and climbing the stairs. The control group received a conventional exercise program which combined aerobic, resistance, flexibility and walking training.

**Outcome measures:** The primary outcome was the pulmonary function test. The secondary outcomes included: a 6-minute walk test with Borg scale (0 to 10) rating of exertion, 10-metre walk test, basic activities of daily living, trunk control test, lower extremity motor score, muscle tone of lower limb, bone mineral density, and distal femoral cartilage at baseline and upon completion of treatment.

**Results:** After 4 weeks of trainings, the experimental group improved more on the forced vital capacity (MD 0.53, 95%CI 0.01 to 1.06), predicted FVC% (MD 19.59, 95%CI 6.63 to 32.54) and forced expiratory volume in 1s (MD 0.61, 95%CI 0.15 to 1.07); BADL (MD 19.75, 95%CI 10.88 to 28.62); and distal femoral cartilage than the control group. Participants completed 6-minute walk test with median 17.3 meters while wearing the exoskeleton. There was no difference in trunk control test, lower extremity motor score, muscle tone, bone mineral density and adverse event.

**Conclusions:** In people with lower thoracic neurological level of spinal cord injury, exoskeleton-assisted walking training instead of a conventional excise program has potential benefits to facilitate pulmonary ventilation function, walking, basic activities of daily living and thickness of cartilage.

**Trial registration:** ChiCTR2000034623

## Introduction

Spinal cord injury (SCI) is a worldwide life-disrupting pathological condition with estimated 17,810 injuries occurred in the United States in 2017,^1,2^ and 3.5 per million in the United Kingdom each year.^3^ Restoring functions (neuromusculoskeletal and respiratory function, etc.), activities of daily living (walk, grasp, lift, etc.), quality of life and life expectancy post-SCI remain to be medical challenges.^4^ Despite great progress in treatments, it is still difficult to achieve the recovery of motor that leads to immobility.^5^ Due to accelerated aging, lifestyle factors, and decreased mobility, even incomplete SCI lesions as low as L4 should be considered as risk factors for cardiopulmonary disability.^6^ A longitudinal decline was reported in forced vital capacity (FVC) and forced expiratory volume in 1 second (FEV_1_),^7^ that indicates persons with SCI are more likely to suffer pulmonary capacity deficit.^8^

Exercise and Sports Science Australia recommended people with SCI to undertake a combined exercise program which contained moderate aerobic exercise (>30min, >5d/week), moderate strength training and flexibility training (>2d/week) for maintaining respiratory function, quality of life and functional independence.^9-12^ However, the improvements are limited for people with SCI due to lower extremity motor lesions and training types.^13^ Exercise of upper body limits their maximal exercise capacity and puts them at a disadvantage compared with leg exercise.^14^ Hence, a new rehabilitation therapy consisting leg exercise is needed for improving the function and independence of people with SCI.

Exoskeleton-assisted walking (EAW) refers to a robotic suit worn on the body enabling a person with paralysis to stand and walk,^15^ that has been confirmed to help individuals with thoracic and lumbar SCI to walk safely.^16-18^ A quantity of studies^19,20^ has manifested the EAW can be used a moderate-intensity level of exercise according to the heart rate, oxygen demand, and rate of perceived exertion. Nevertheless, these clinical trials were designed as observational study, and the changes of pulmonary ventilation function followed by EAW have not been reported yet. It is unclear that whether better pulmonary caused by EAW training performed better exercise capacity in walking. There is lack of evidence that related to the effect of EAW training on respiratory function, quality of life and functional independence for people with SCI. Further, Further, the relation between musculoskeletal function, pulmonary function, and walk ability are not yet completely understood.

Therefore, the research question for this randomised trial was:

In people with SCI, do EAW training provide equivalent benefits on the pulmonary ventilation and musculoskeletal function, and the functional independence to those obtained in a conventional exercise program?

## METHODS

### Design

This was a single-blinded, randomized controlled efficacy trial with 2 parallel groups and intention-to-treat analysis. The study protocol has been registered at Chinese Clinical Trial Registry (ChiCTR2000034623) and approved by the medical ethics committee of West China Hospital of Sichuan University. All patients were informed of the procedure, the use of their data and images for research. They understood the purposes and provided written informed consent according to the 1964 Declaration of Helsinki prior to their participation.

The individuals were randomly divided into one of two groups in a 1:1 ratio by simple randomization method, using a computer-generated simple random table. The sequences were preserved using closed envelop method by one researcher who did not participant in the trainings and assessments. Clinical data except walking parameters were measured at baseline and post-training by two clinical researchers who were blinded and did not know the group to which each participant belonged. Walking parameters were assessed by two researchers who participated in an EAW program or a conventional program. Clinical data was recorded after averaging.

### Participants

From July 2020 to March 2021, we prospectively enrolled all individuals aged between 15 and 65 years with a diagnosed SCI between T4 and L1 at least 1 month. Participants were recruited from inpatients in 3 units of Rehabilitation centre, West China Hospital, Sichuan University. In addition, the eligible individuals met the following inclusion criteria: (1) American spinal injuries association impairment scale (AIS)^21^ classified with A, B or C, (2) the height was between 1.50 meters and 1.85 meters and (3) stopped smoking for over 6 months. Exclusion criteria were: (1) spasticity of any lower extremity muscle scored over 2 according to the Modified Ashworth Scale,^22^ (2) unstable fracture, (3) had EAW training experience before, (4) hypertension, (5) severe osteoporosis (bone mineral density t-score< -3.5), (6) any respiratory or other neurological diseases, (7) overweight (> 100 Kg).

### Interventions

All participants were received 16 sessions of exercise training for 50 to 60 minutes per session, 1 session per day, 4 sessions per week for 4 weeks. Exercise intensity was requested to reach 60% to 70% maximal heart rate (HR, HR_max_=220 - age) that is checked with the values of a heart rate sensor (Polar H10, POLAR^®^ China). All participants were scheduled to perform occupational therapy, endurance training cycling using upper limb, and biofeedback therapy once per day. Medications and rehabilitation nursing were ordered based on the medical condition.

#### Experimental group

The AIDER (AssItive DEvice for paRalyzed patient) powered robotic exoskeleton (generation IV, Buffalo Robot Technology Co. Ltd, Chengdu, China) was used for the EAW training. A mobile phone application (Android) as an additional treatment system switched different training modes (from sit to stand, walk, climb, etc.). Body-weight-supported AIDER system was applied at the beginning of this program to protect users. All subjects were individually fitted to the robotic exoskeleton according to pelvic width, thigh length, and shank length. Training session included sitting, standing, walking, climbing stairs and slope with maximal assistance-walking mode under monitoring of experimented therapists. A progressive EAW program was designed for participants that shown in Table 1.

**Table 1.** Description of the EAW program in the experimental group

| Exercise | Session |
| --- | --- |
| From sit to stand and standing training using body-weight-supported AIDER system, controlled by therapist. | 1 |
| Standing training and shifting of the body centre of gravity using body-weight-supported AIDER system, controlled by patient. | 1 |
| Walking training using body-weight-supported AIDER system, controlled by patient. | 1 |
| From sit to stand, standing training and shifting of the body centre of gravity using AIDER system, controlled by patient. | 2 |
| Walking training (walk straight) using AIDER system, controlled by patient. Turning around with the help of therapists. | 1 |
| Walking training (walk straight and turn around) using AIDER system, controlled by patient. | 6 |
| Climb the slope using AIDER system, controlled by therapist. | 1 |
| Climb the slope using AIDER system, controlled by patient. | 1 |
| Take the stairs using AIDER system, controlled by therapist. | 1 |
| Take the stairs using AIDER system, controlled by patient. | 1 |

EAW = exoskeleton-assisted walking, AIDER = Asslitive DEvice for paRalyzed patient

#### Control group

Individuals in the control group undertook a conventional exercise program that included strength training using dumbbell between 5 kg and 20 kg, dynamic balance training in sitting or standing position, flexibility training, and walking training with brace. The time proportion of each training depended on the situations of individuals.

### Outcome measures

Outcome measures were collected and analysed at the baseline and end of 16-session intervention period.

#### Primary outcome

Pulmonary ventilation function test was completed with a computerized spirometer (Vyntus™ SPIRO PC Spirometer, Vyaire Medical Inc., Mettawa, US) based on the standardized procedures as the American Thoracic Society^23^ described. To determine pulmonary ventilation function, participants performed test seating in the wheelchair and were forbidden to disclose their intervention assignment to the assessor. The test was performed with the participants wearing a nose clip. If the participant coughed or made a mistake, the numerical values were not recorded. Three repeated maneuvers were performed, separated by a five-minute rest and the best result was recorded automatically. The test consisted of the assessments of FVC, predicted FVC%, FEV_1_, forced expiratory flow (FEF_25/50/75_), peak expiratory flow (PEF), and maximal voluntary ventilation (MVV).^24^

#### Secondary outcomes

Walking parameters were assessed by the 6MWT (6-minute walk test) for distance and 10MWT (10-metre walk test) for speed. Tests were performed in door in accordance with the guidelines of the American Thoracic Society^25^. The participant’s heart rate, and the rate of perceived exertion (RPE) based on the Borg scale^26^ during the 6MWT were recorded. Participants in EAW group were allowed to wear the exoskeleton, while those in control group using the knee-ankle-foot orthoses if they had one. Modified Barthel index (MBI) were reported to demonstrate the ability of basic activities of daily living (BADL).^27^

Moreover, trunk control test (TCT)^28^ and lower extremity motor score (LEMS)^29^ were reported to clarify the reasons of ventilation and walking improvement, and the recovery of muscle strength. The thickness of distal femoral cartilage that consisted of medial condyle, intercondylar area, and lateral condyle measured by musculoskeletal ultrasound were compared. The value of Wards triangle bone mineral density (BMD) evaluated by dual-energy X-Ray absorptiometry, and muscle tone of lower limb estimated by modified Ashworth scale (MAS)^30^ were recorded. The rate of adverse events was used for safety indicator.

### Data Analysis

As for the sample size, a pilot study^31^ was performed for determinizing the effect size (1.03975) of primary outcome (FVC). It was calculated that at least 16 participants were required for each group according to a significance level of 5% and study power of 80%. The final sample size was 40 participants when considered the rate of dropping out as 20%.

Analyses were completed according to intention-to-treat principle. Data with normally distributed were recorded as means (SDs), others were described as median (IQR) where necessary. The differences between groups were reported as means (95% CI). Pearson correlation test or Spearman rank correlation test was performed to discuss the relation between the results of 6MWT, TCT and ventilation parameters with statistically significant difference between groups.

## RESULTS

### Flow of participants

A total of 134 individuals with SCI were screened from July 2020 to March 2021, of which 94 were excluded as per exclusion criteria (n=81) or declining to commit the full participation (n=13). 40 eligible individuals were randomized to either EAW group (n=20) or control group (n=20). Two groups were comparable on the baseline characteristics (Table 2). Figure 1 represents the Consolidated Standards of Reporting Trials (CONSORT) diagram.

**Table 2.** Baseline characteristics of participants

| Characteristic | EAW (n=20) | Con (n=20) |
| --- | --- | --- |
| Demographic |  |  |
| Age (yr), mean (SD) | 36.6 (11.7) | 37.7 (12.6) |
| Height (cm), mean (SD) | 166.2 (6.4) | 170.0 (6.2) |
| Weight (kg), mean (SD) | 56.1 (9.1) | 64.0 (10.0) |
| Gender, No. (%) |  |  |
| Female | 4 (20.0) | 3 (15.0) |
| Male | 16 (80.0) | 17 (75.0) |
| Clinical |  |  |
| NLI, No. (%) |  |  |
| T4-T10 | 12 (60.0) | 12 (60.0) |
| T11-L1 | 8 (40.0) | 8 (40.0) |
| AIS, No. (%) |  |  |
| A | 15 (75.0) | 10 (50.0) |
| B | 3 (15.0) | 2 (10.0) |
| C | 2 (10.0) | 8 (40.0) |
| DOI (mth), median (IQR) | 2.0 (4.5) | 2.0 (0.5) |

EAW = exoskeleton-assisted walking group, Con = control group, NLI = Neurological level of injury, AIS = American spinal injuries association impairment scale, DOI = duration of injury

**Figure 1.**
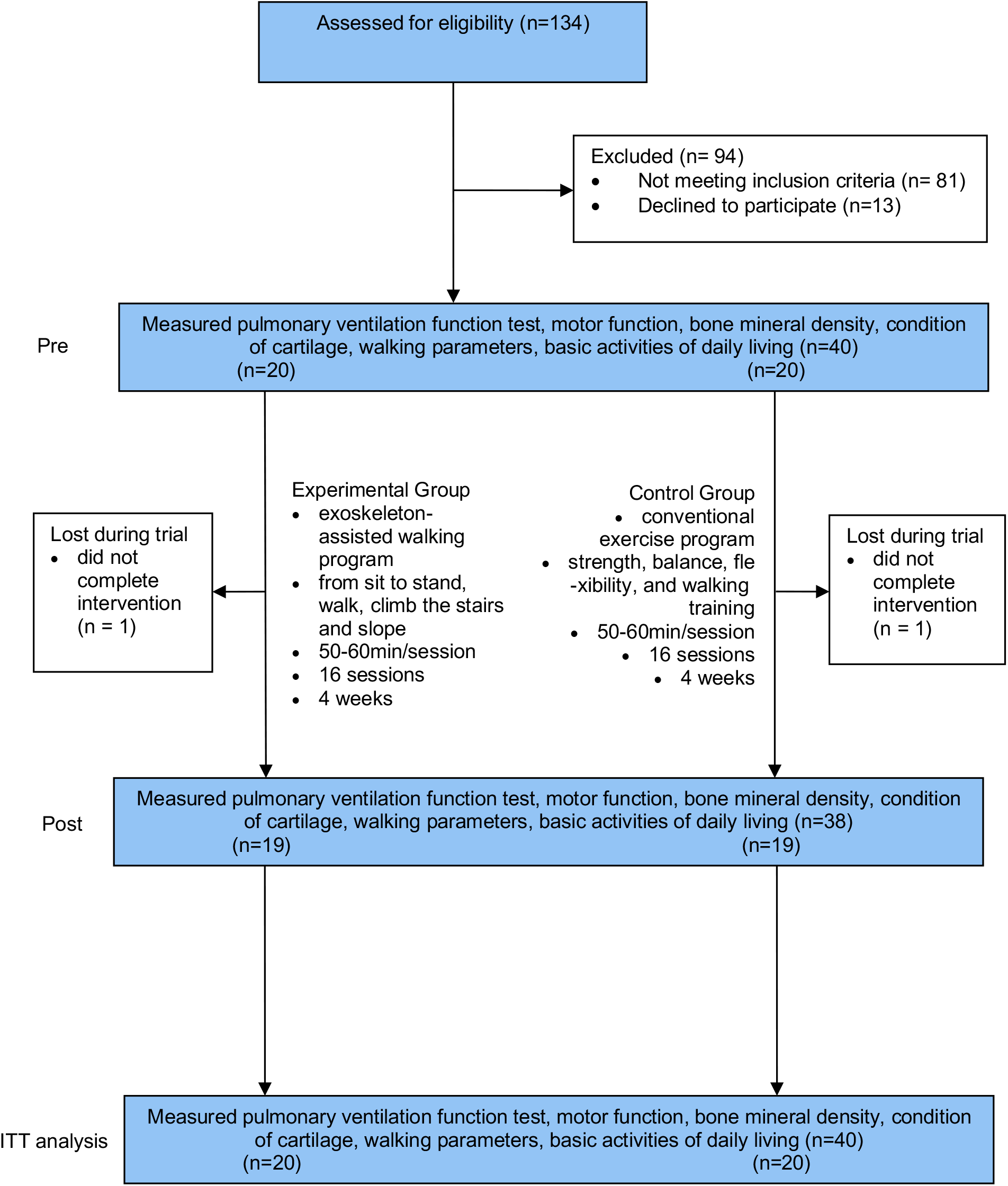
Design and flow of participants through the trial.

### Compliance with the study protocol

After pre-intervention assessment, one individual in the EAW group, and one in the control group withdrew the study and did not accept the final assessment because of own desire to be discharged from hospital.

### Effect of intervention on pulmonary ventilation function

EAW group showed improvements in pulmonary ventilation function by the end of 16-session intervention period. EAW training was estimated to be more favourable than conventional exercise program for several measures of pulmonary ventilation function, including FVC, FVC%_predictied_, and FEV_1_. The results for FEF_25-75_, PEF, and MVV supported that EAW training might be equivalent to conventional exercise program. The estimates and their 95% confidence intervals are presented in Table 3.

**Table 3.**
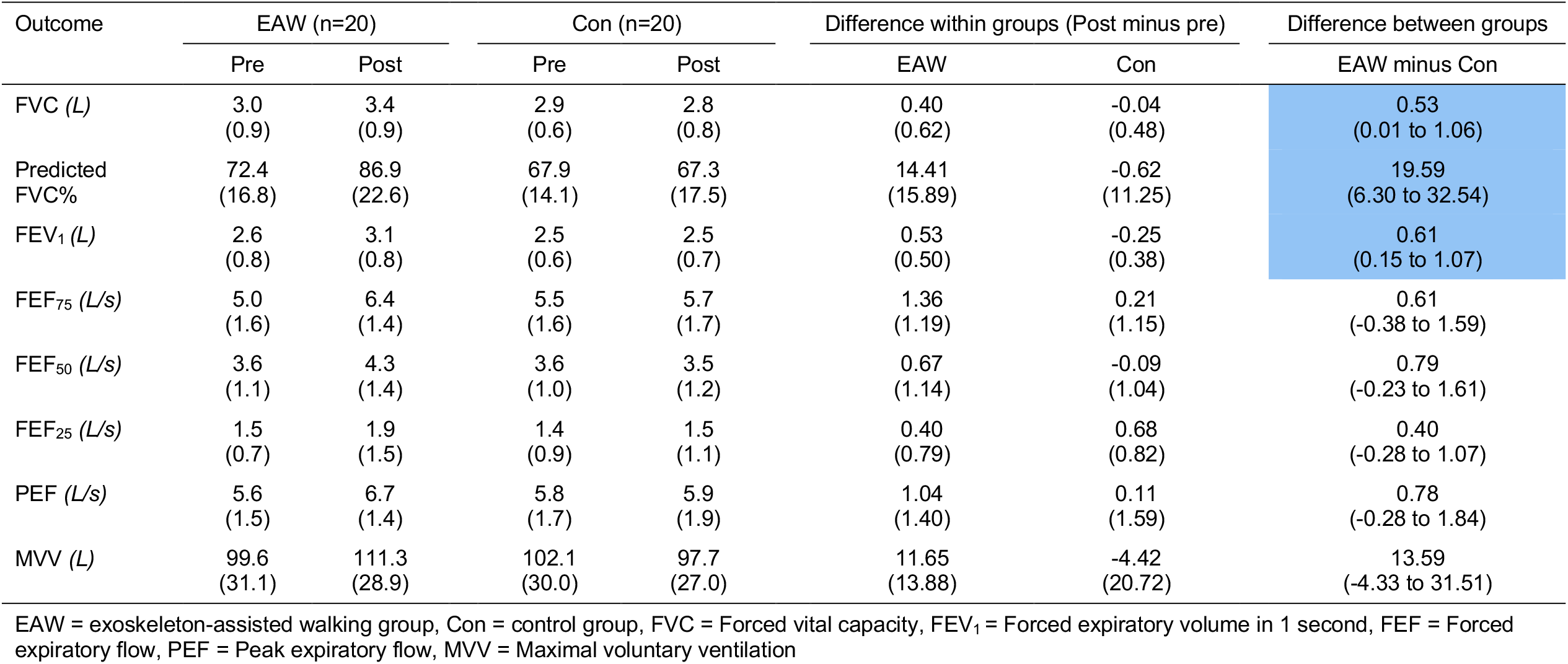
Mean (SD) of groups, mean (SD) difference within groups, and mean (95% CI) difference between groups for the pulmonary ventilation function

### Effect of intervention on walking parameters and BADL

Of the 21 participants who completed the final 6MWT and 10MWT, 2 were in the control group, 19 in the EAW group while wearing the exoskeleton in door. The distance and speed in the EAW group were 15.9 (5.0) meters and 0.049 (0.164) m/s, respectively. PRE and heart rate were 3.47 (1.47) and 114.5 (18.0) bpm during the 6MWT.

Both groups showed statistical improvement in the BADL after interventions. The improvement was estimated to be 19.75 greater in the EAW group (95% CI 10.88 to 28.62) than in the control group. These better results on the MBI were achieved with a mean difference of 28.8 (SD 15.0) within the EAW group and 14.0 (SD 11.5) within the control group by the end of trainings. These results are presented in Table 4.

**Table 4.** Mean (SD) or median (IQR) of groups, mean (SD) difference within groups, and mean (95% CI) difference between groups for motor function, the distal femoral cartilage and bone mineral density (BMD)

| Outcome | EAW (n=20) |  | Con (n=20) |  | Difference within groups (Post minus pre) |  | Difference between groups |
| --- | --- | --- | --- | --- | --- | --- | --- |
|  | Pre | Post | Pre | Post | EAW | Con | EAW minus Con |
| TCT | 54.4<br>(23.6)<sup>†</sup> | 74.0<br>(35.8)<sup>†</sup> | 40.6<br>(19.6) | 61.0<br>(26.0) | 25.0<br>(35.0)<sup>†</sup> | 19.6<br>(19.2) | N/A |
| MAS, n (%) |  |  |  |  |  |  |  |
| Grade 0 | 18 (90) | 18 (90) | 18 (90) | 18 (90) | N/A | N/A | N/A |
| Grade 1 | 1 (5) | 1 (5) | 2 (10) | 1 (5) | N/A | N/A | N/A |
| Grade 1+ | 1 (5) | 1 (5) | 0 (0) | 1 (5) | N/A | N/A | N/A |
| LEMS<sup>†</sup> | 1.0<br>(10.8) | 1.5<br>(7.8) | 0<br>(7.5) | 5<br>(13.8) | 0<br>(0) | 0.5<br>(4.5) | N/A |
| Left medial condyle<br>(mm) | 1.49<br>(0.51) | 1.80<br>(0.31) | 1.59<br>(0.54) | 1.46<br>(0.52) | 0.31<br>(0.50) | -0.13<br>(0.35) | 0.34<br>(0.06 to 0.61) |
| Left intercondylar area<br>(mm) | 2.02<br>(0.25) | 2.31<br>(0.45) | 1.94<br>(0.50) | 1.85<br>(0.44) | 0.29<br>(0.41) | -0.09<br>(0.45) | 0.47<br>(0.18 to 0.75) |
| Left lateral condyle<br>(mm) | 1.99<br>(0.38) | 2.13<br>(0.30) | 1.91<br>(0.44) | 1.87<br>(0.43) | 0.14<br>(0.35) | -0.04<br>(0.28) | 0.26<br>(0.02 to 0.50) |
| Right medial condyle<br>(mm) | 1.52<br>(0.40) | 1.78<br>(0.28) | 1.55<br>(0.42) | 1.52<br>(0.39) | 0.26<br>(0.34) | -0.03<br>(0.38) | 0.26<br>(0.04 to 0.47) |
| Right intercondylar area<br>(mm) | 2.04<br>(0.48) | 2.30<br>(0.39) | 1.85<br>(0.50) | 1.94<br>(0.48) | 0.26<br>(0.34) | -0.09<br>(0.40) | 0.32<br>(0.03 to 0.60) |
| Right lateral condyle<br>(mm) | 1.97<br>(0.43) | 2.20<br>(0.28) | 2.05<br>(0.54) | 1.90<br>(0.51) | 0.24<br>(0.45) | -0.15<br>(0.39) | 0.31<br>(0.04 to 0.60) |
| BMD<br>(g/cm<sup>3</sup>) | 0.78<br>(0.14) | 0.81<br>(0.20) | 0.90<br>(0.19) | 0.89<br>(0.22) | 0.04<br>(0.09) | -0.02<br>(0.13) | 0.07<br>(-0.21 to 0.06) |

EAW = exoskeleton-assisted walking group, Con = control group, TCT = trunk control test, MAS = modified Ashworth scale, LEMS = lower extremity motor score, BMD= bone mineral density, <sup>†</sup>= median (IQR)

### Effect of intervention on motor function

Both groups showed statistical improvement in the TCT by the end of trainings, while the improvement of the EAW training might be equivalent to conventional exercise program. Additionally, the control group showed improvement in LEMS by the end of trainings. Nonetheless, the results of LEMS and MAS did not have estimates that clearly favoured one intervention over the other. These results are presented in Table 4, with individual-participant data in Table 6 on the eAddenda.

**Table 5.** Mean (SD) of groups, mean (SD) difference within groups, and mean (95% CI) difference between groups for the basic activities of daily living (BADL)

| Outcome | EAW (n=20) |  | Con (n=20) |  | Difference within groups<br>(Post minus pre) |  | Difference<br>between groups |
| --- | --- | --- | --- | --- | --- | --- | --- |
|  | Pre | Post | Pre | Post | EAW | Con | EAW minus Con |
| MBI | 34.8<br>(14.7) | 65.0<br>(17.5) | 29.8<br>(14.7) | 42.5<br>(15.0) | 28.8<br>(15.0) | 14.0<br>(11.5) | 19.8<br>(10.9 to 28.6) |

EAW = exoskeleton-assisted walking group, Con = control group, MBI = modified Barthel index

**Table 6.** Outcomes of correlation of physical function and fitness

| Characteristic | Correlation coefficient | p-value |
| --- | --- | --- |
| FVC – distance | 0.122 | 0.620 |
| Predicted FVC% – distance | 0.540 | 0.017 |
| FEV<sub>1</sub> – distance | 0.075 | 0.761 |
| TCT – distance | 0.427 | 0.068 |
| PRE – distance | -0.605 | 0.006 |
| FVC – TCT | -0.062 | 0.801 |
| Predicted FVC% – TCT | 0.210 | 0.387 |
| FEV<sub>1</sub> – TCT | 0.044 | 0.858 |
| RPE – TCT | -0.394 | 0.095 |
| FVC – RPE | -0.295 | 0.220 |
| Predicted FVC% – RPE | -0.596 | 0.007 |
| FEV<sub>1</sub> – RPE | -0.290 | 0.228 |

FVC = Forced vital capacity, FEV<sub>1</sub> = Forced expiratory volume in 1 second, TCT = trunk control test, RPE = rate of perceived exertion

### Effect of intervention on bone and cartilage

EAW training was estimated to be more favourable than conventional exercise program for protecting distal femoral cartilage. The effect on BMD was unclear (MD 0.07 g/cm^3^, 95% CI –0.21 to 0.06). The confidence intervals around these effect estimates confirmed the benefit (Table 5).

The exercise training in both groups was well tolerated and there were 3 adverse events in the EAW group. Adverse events consisted of fall (n=1) and heel indentation (n=2), that disappeared within 24 hours and caused no injury to participants. Individual participant data for all outcomes are presented in Table 6 on the eAddenda.

## DISCUSSION

For individuals with T4 to L1 SCI, EAW training instead of conventional exercise program improved the pulmonary ventilation function, trunk control, and BADL. Moreover, it promoted the walking ability, and protected the cartilage. However, it is estimated that neither were effective for motor function recovery and bone loss prevention. It is essential to consider whether the benefits from EAW training are robust (as reflected in the confidence intervals) to warrant recommending it over conventional exercise program.

The estimate of the differences between the groups on the predicted FVC% and FEV_1_ (19.6% and 0.61L) were more beneficial than the minimal clinically important difference. Because the minimal clinically important difference for predicted FVC% and FEV_1_ are 2-6% by distribution-based method,^32^ and 0.1L according to previous study by the anchor-based method.^33^ Moreover, the estimate of the effects on predicted FVC% from post- to pre-intervention in EAW group and control group and were 14.4% and -0.6%, respectively. And effects on changes of FEV_1_ were 0.53L and -0.02L for EAW trainings and conventional exercise program.

Both groups were required equal intensity and amounts of time training in this study. Therefore, the effects on pulmonary ventilation function might be considered as a ‘potential’ bonus, that was meaningful for people with SCI. Upper limb exercise are limited for improving a maximal external power output, peak oxygen consumption, heart rate, total peripheral resistance, and responses for cardiac stroke volume.^14^ Another underlying mechanism might be the potential of EAW to recruit trunk muscles.^34,35^ Alamro et al. ^36^ and Guan et al.^37^ have demonstrated that overground walking by exoskeleton elicits greater activation of trunk muscles compared to treadmill walking, even after controlling for the use of hand-held assistive devices. The benefits for pulmonary ventilation function are consistent with other research,^20,38-40^ although it reported a higher VO_2 peak_ among the incomplete SCI individuals after EAW training.

Robotic exoskeleton was achievable to enhance the walking capacity. The greater distance on the 6MWT was achieved with a higher predicted FVC% (r=0.54) and less exertion (r=-0.61). The improvement of walking performance was not caused by the recovery of lower limb motor function. All participants were able to perform walking while wearing the exoskeleton, that was similar to McIntosh et al.^41^ and Sale et al.^42^ Longer training duration may result in longer distance. Benson et al.^43^ found the minimal distance of 6MWT after 10-week trainings was 91 meters which was more than 5 times than our average distance.

Additionally, we found that individuals with lower neurologic injury level (T11 to L1) had similar mean improvement (15.6 meters) than others (16.0 meters). The age or neurologic injury level did not perform as an associated factor for walking distance. Participants who younger than 40 years-old completed 15.5 meters during 6WMT, with that of 17.3 meters for older participants. This was inconsistent with previous study^44^ that reported walking speed and performances were significantly associated with injury level. The inconsistence caused by product update according to the result of Guanziroli et al.^45^

Beyond this, the application of EAW and product update led to the differences between groups on walking and going stair, that two issues of MBI. Hence, participants in the EAW group were accessible to gain the scores on these two aspects with exoskeleton. However, besides EAW and conventional exercise program, occupational therapy is also benefit for the ability of ADL.^46^ Thus, the possible effect of two technologies on BADL is overestimated and needs to be considered.

In addition, the effects on bone and cartilage were further explored. There was an increase of BDM in the EAW group, although the estimate was unclear and the values before intervention were different. The effect on EAW on BMD of Wards triangle was consistent with the study of Karelis et al.^47^, which reported an improvement with clinical meaning measured by peripheral quantitative computed tomography on tibia. Different training prescriptions lead to different effects on joint, although the effect on cartilage was merely reported in EAW training. Optional duration (less than 1 hour) is helpful for increasing the thickness of distal femoral cartilage according to Yilmaz et al.,^48^ while higher intensity is harmful for cartilage.^49^ Therefore, the EAW had potential benefit for protecting the cartilage for SCI and the training prescription in this study was suitable.

This study was limited to the number of training session, although our feasibility study has proved this training period realized the application of EAW and was most achievable in inpatient rehabilitation in the health care system. Although we tried to avoid the detection bias, it was inevitable during the 6MWT. In this study, the range of age is 40 and the used equation underestimates the HR_max_ in older adults.^50^ Moreover, the validity of equation has not been established in a study sample that included an adequate number of individuals with SCI. This might have resulted in misestimation of the training intensity. Further, gait parameters should be recorded and compared while walking without the exoskeleton.

In summary, the study successfully manifested that EAW training has the potential to improve performance in pulmonary ventilation function, trunk control, and BADL among individuals with SCI at least equivalently with conventional exercise program. Additionally, it promoted the walking ability, and protected the cartilage better than conventional exercise program. However, it is estimated that neither were effective for motor function recovery and bone loss prevention.

### What was already known on this topic

Pulmonary ventilation and motor function are most frequently affected by spinal cord injury. Recovery of motor function is hard to achieve, while progression in pulmonary ventilation function helps to improve motor performance and activities of daily livings. Conventional exercise programs help to maintain pulmonary ventilation function and activities of daily livings. Exoskeleton-assisted walking is a new technique for gain walking ability among people with spinal cord injury.

### What this study adds

In people with spinal cord injury, exoskeleton-assisted walking training improve performance in pulmonary ventilation function, trunk control, and basic activities of daily livings among individuals with SCI equivalently with conventional exercise program. Other benefits of the exoskeleton-assisted walking were better improvement in walking ability and protection for joint cartilage. However, neither were effective for motor function recovery and bone loss prevention.

### Ethics approval and consent to participate

This study received approval from the Ethics Committee of West China Hospital of Sichuan University and registered at the Chinese Clinical Trial Registry with the following identifier: ChiCTR2000034623. Participants gave written informed consent before starting data collection.

## Data Availability

All data produced in the present study are available upon reasonable request to the authors

## Acknowledgments

The authors are grateful for the assistance from Professor Hong Cheng. Additionally, we also thank the participants who were involved in the project.

## Availability of data and materials

The dataset used in the study is available from the corresponding author on reasonable request.

